# Clinical and laboratory evaluation of patients with SARS-CoV-2 pneumonia treated with high-titer convalescent plasma: a prospective study

**DOI:** 10.1101/2020.07.20.20156398

**Authors:** Michele L Donato, Steven Park, Melissa Baker, Robert Korngold, Alison Morawski, Xue Geng, Ming Tan, Andrew Ip, Stuart Goldberg, Scott Rowley, Kar Chow, Emily Brown, Joshua Zenreich, Phyllis McKiernan, Kathryn Buttner, Anna Ullrich, Laura Long, Rena Feinman, Andrea Rincourt, Marlo Kemp, Mariefel Vendivil, Hyung Suh, Bindu Balani, Cristina Cicogna, Rani Sebti, Abdulla Al-Khan, Steven Sperber, Samit Desai, Stacey Fanning, Danit Arad, Ronaldo Go, Elizabeth Tam, Keith Rose, Sean Sadikot, David Siegel, Martin Gutierrez, Tatyana Feldman, Andre Goy, Andrew Pecora, Noa Biran, Lori Leslie, Alfred Gillio, Sarah Timmapuri, Michele Boonstra, Sam Singer, Sukhdeep Kaur, Ernest Richards, David S Perlin

## Abstract

**OBJECTIVES:** To determine the rate of intubation, overall survival, viral clearance, and the development of endogenous antibodies in patients with COVID-19 pneumonia treated with convalescent plasma containing high levels of neutralizing anti-SARS-CoV-2 antibodies. We also aimed to describe the laboratory parameters of the plasma products.

**DESIGN:** This was a phase IIa, single institution, prospective study in adults hospitalized with SARS-CoV-2 pneumonia.

**SETTING:** Hackensack University Medical Center, a 770-bed research and teaching hospital located in Bergen County NJ, 11 km from New York City. The study was conducted between April 15 and June 18, 2020.

**PARTICIPANTS:** 47 hospitalized adult patients were treated: 32 in the non-mechanically ventilated group and 15 in the mechanically ventilated group. All patients had confirmed SARS-CoV-2 pneumonia by radiographic and laboratory evaluation.

**INTERVENTION:** Fresh or frozen convalescent plasma from donors with high titers of viral neutralizing antibodies was administered.

**MAIN OUTCOME MEASURES:** Incidence of intubation, overall survival, and discharge rate of patients divided in cohorts based on severity of disease. Description of infused plasma characteristics. Evaluation of recipients’ pre-treatment viral immunity, immunity transfer from convalescent plasma administration, and late immunity 30 and 60 days after treatment. Rates of viral clearance by nasopharyngeal PCR at 10 and 30 days. Outcomes of patients with no pre-treatment immunity. Survival comparison with institutional data for each cohort.

**RESULTS:** Analysis for the non-mechanically ventilated patients showed an intubation rate of 15.6% (95% CI: 5.3%-32.8%) and a day-30 survival rate of 87.5% (28/32; 95% CI:70.2%-96.4%). The overall survival for a comparative group based on institutional data was 66% (675/1023; p=0.012). The rates of negative nasopharyngeal swab by PR-PCR on day+10 and +30 post treatment were 42.9% (95% CI: 24%-63%) and 78% (95% CI: 56%-93%) respectively. Patients mechanically ventilated had a day-30 mortality of 46.7% (95% CI:21.3%-73.4%); the mortality for a comparative group based on institutional data was 68.5% (217/317; p=0.093). The rates of negative nasopharyngeal swab by PR-PCR at day+10 and +30 was 85.7% (95% CI: 42-100%; n=7) and 100% (95% CI: 63-100%; n=8). Seven patients (15%) had no pre-infusion immunity, and all were found to have anti-SARS-CoV-2 neutralizing titers three days post infusion. All evaluable patients were found to have neutralizing antibodies on day+30 (n=30) and on day+60 (n=12) post treatment. There was no difference in outcomes within the ranges of high antiviral neutralizing titers used, mostly greater than 1:1000. There was also no difference between fresh or frozen plasma. The only adverse event was a mild rash in one patient.

**CONCLUSION:** In this study of adult patients hospitalized with SARS-CoV-2 pneumonia, convalescent plasma was safe and conferred effective transfer of immunity while preserving endogenous immune response. Intubation rates, survival rates compared with institutional data, and viral clearance rates, support the continued evaluation of this antiviral modality.

**STUDY REGISTRATION:** ClinicalTrials.gov NTC04343755

## Introduction

As of July 11, 2020, over twelve million people around the world have been infected with the severe acute respiratory syndrome coronavirus 2 (SARS-CoV-2) and over 550,000 have died.^1^ The human and economic impact, unprecedented in our generation, has mobilized the medical community in search of effective treatment strategies. The angiotensin-converting enzyme 2 (ACE2) is necessary for SARS-CoV-2 to enter human cells.^2^ The initial phase of the disease occurs when SARS-CoV-2 infects the respiratory epithelial cells, but in addition to lung tissue, expression of ACE2 in found broadly, including in renal, intestinal, and adipose cells, leading to a wide viral impact on the host.^3^ Moreover, ACE2 has been shown to be upregulated by SARS-CoV-2 infections.^4^ The innate immune response to the viral infection leads to the release of cytokines, and the ensuing cytokine storm results in acute respiratory distress syndrome and multiorgan failure.^5^ The natural response to viral infections including coronaviruses, is the production of high affinity immunoglobulin G (IgG) during the adaptive immune response.^6^ SARS-CoV-2 has been associated with the suppression of this T cell-mediated immune response, which bring into question the quality of the adaptive immunity in severely ill patients.^7^ A therapeutic intervention focused on viral neutralization is therefore a priority.

Convalescent plasma as a method of passive immunity transfer has a long history dating to the Spanish flu of 1918.^8^ More recently, convalescent plasma was deployed in the management of SARS^9^ and MERS^10^, with evidence of viral neutralization. Convalescent plasma therapy in the setting of SARS-CoV-2 infection is currently an active field of investigation^11-23^, but information on immune transfer, the subsequent endogenous response, and the clinical course of patients at different stages of the disease remains incomplete. Furthermore, since the development of neutralizing antibody titers varies among COVID-19 recovered patients, convalescent plasma is a heterogeneous product of varying potency. In this study, we investigated both the clinical and laboratory parameters characterizing patients treated with high-titer anti-SARS-CoV-2 neutralizing convalescent plasma.

#### WHAT IS ALREADY KNOWN ON THIS TOPIC

In March 2020, the first report of 5 patients with severe COVID-19 disease treated with convalescent plasma was published.^11^ This initial report, and previous data on the use of convalescent plasma for the management of other viral infections, renewed interest in this approach. As of July 11, 2020, 16 publications reporting on the clinical outcome of patients receiving convalescent plasma have been published, excluding single case reports. The largest study determined the safety of this modality,^12^ and a single randomized study reported on the time to improvement of symptoms.^13^

#### WHAT THIS STUDY ADDS

Novel aspects of our study include the comparison of fresh and frozen plasma, the evaluation of immunoglobulin subset dosing and the plasma antiviral titer levels. Importantly, we also evaluated the effectiveness of antiviral immunity transfer and the late impact on the recipients’ antiviral immunity. We compared the survival of treated patients to our institutional data for similar patient cohorts. We also analyzed the outcome parameters of patients with no or minimal pre-treatment immunity. Strengths of this study include the prospective and complete collection of clinical and basic science information on recipients and plasma components. This study offers the most comprehensive analysis aiming at understanding the clinical and laboratory impact of this therapeutic approach. Clinicians will be able to apply this knowledge when managing patients with COVID-19 disease. Furthermore, this information will assist in the design of future research.

## Methods

### Study design

We conducted a single institution prospective phase IIa clinical trial. The study was performed at Hackensack University Medical Center, a tertiary medical center and home to the John Theurer Cancer Center. Patients were included if they were aged 18 years or older and were hospitalized for the management of symptoms associated with a documented infection with SARS-CoV-2. Patients were excluded for a history of severe transfusion reactions, infusion of immunoglobulins with 30 days, AST or ALT greater than 10 times the upper limit of normal, requirement for vasopressors and dialysis. Patients requiring intermittent vasopressors for sedation management were treated. Prospective plasma donors were included if they were aged 18 to 60 years, had a history of a positive nasopharyngeal swab for COVID-19 or a positive antibody test, were at least 14 days from resolution of symptoms, had one subsequent negative swab, were found to have high titers of neutralizing antibodies against SARS-CoV-2 (>1:500), and met institutional and FDA regulations for donation of blood products.

### Procedures

Volunteer donors were recruited through advertising in the local community. Individuals who agreed to participate and gave informed consent were evaluated at the John Theurer Cancer Center where they underwent a physical examination, completed a donor health questionnaire, had a nasopharyngeal swab for SARS-CoV-2 and blood drawn for complete blood count and chemistry, infectious disease markers, and HLA antibodies for female donors.

The presence of SARS-CoV-2 neutralizing antibodies was evaluated using the previously described COVID-19 ELISA protocol with recombinant spike receptor binding domain (RBD) as capture antigen.^24^ High-titer sera was evaluated for virus neutralization in a viral cytopathic assay performed with Vero E6 cells at 100 × the TCID50 value. The assay using SARS-CoV-2 in Vero E6 cells was established under biosafety level 3 (BSL-3) containment to assess intracellular inhibitory potencies of small molecules. Final assay conditions were 30,000 Vero E6 cells per well and virus at a MOI of 0.01-0.05 in 200 ul. The plates were incubated for 48 or 72 hours at 37°C and 5% CO_2_. Viral ToxGlo™ Luminescent Cell Viability Kit (Promega Corp, Madison, WI, USA) was used to provide a semi-quantitative measure of virus infected cell viability. To assess the distribution of the different antibody isotypes/subclasses in the plasma samples, another ELISA was performed with different secondary antibodies. Donors found to have neutralizing IgG Spike RBD greater than 1:500 were selected for plasma donation, with a preference for titers 1:1000-10,000 and >1:10,000. Donors underwent plasmapheresis using the Trima Accel® system for either a planned fresh infusion of 500 mL or for cryopreservation in aliquots of 200 mL.

Recipients were referred by the clinical teams through the institutional COVID-19 research request process and were treated if eligible. A single infusion of convalescent plasma was administered at a rate less than 250 mL per hour. Premedication with diphenhydramine 25 mg IV and hydrocortisone 100 mg IV with or without acetaminophen was given. The use of fresh versus frozen plasma was based solely on the availability of product at the time of request. Exploratory blood work including serology for anti-SARS CoV-2 titers was performed immediately pre-infusion and on day+3, +10, +30 and +60 post treatment. SARS CoV-2 testing by RT-PCR from nasopharyngeal or endotracheal tube secretions was done on day+10 and if positive again on day+30. A 10 mL sample of plasma was collected at the bedside from the donor plasma bag immediately pre-infusion for analysis.

For comparison, we evaluated the outcomes of patients treated for COVID-19 within our hospital system, Hackensack Meridian Health. Data was collected from the electronic health records of patients hospitalized. Patients in the database were selected if SARS-CoV-2 polymerase chain reaction tests were positive. The data was manually abstracted by research nurses and physicians from the John Theurer Cancer Center. We selected patients from this database with characteristics closest to the patients in our two treatment cohorts. For the first cohort we included patients with radiographic evidence of pneumonia, receiving oxygen other than positive pressure ventilation and for the second cohort, patients in ICU receiving invasive or non-invasive positive pressure ventilation.

### Outcomes

The primary endpoint for patients hospitalized for SARS-CoV-2 infection but not receiving invasive or non-invasive positive pressure mechanical ventilation, was to evaluate the efficacy of convalescent plasma in reducing the rate of intubation. The primary objective for patients already receiving invasive or non-invasive positive pressure ventilation was to evaluate the efficacy of convalescent plasma in reducing the mortality rate at day +30. The safety of convalescent plasma was also a primary objective. Secondary objectives for both groups included duration of hospitalization, overall survival, rate of virologic clearance by nasopharyngeal swab RT-PCR at day +10 and +30, impact of donor neutralizing antibody titer levels on the primary objectives, and recipient anti-SARS-CoV-2 titer levels pre-infusion and on days +3, +10, +30 and +60.

### Statistical analysis

It is important to note that at the time of the study’s statistical design in late-March 2020, the availability of outcomes data was more limited. For research purposes across studies, patients with COVID-19 at our institution were divided in three tracks based on acuity, track 1 being attributed to outpatients, track 2 for patients hospitalized but not requiring positive pressure mechanical ventilation, and track 3 for patients receiving positive pressure mechanical ventilation. Our statistical plan for this study included only patients ascribed to tracks 2 and 3. We used a multistage design based on the sequential conditional probability ratio test.^25^ The statistical design was based on the following hypothesis: for track 2 the null hypothesis assumed an intubation rate of 30% and the alternative hypothesis was an intubation rate ≤ 15%. For track 3 the null hypothesis assumed a mortality rate of at least 49% with an alternative hypothesis of ≤ 25%. The design for each track had a type I error rate of 0.1 with statistical power of at least 0.8. The decision to accept or reject the null hypothesis was made based on interim data analysis in a three-stage process. Descriptive statistics were used to characterize the baseline profile of the subjects and exploratory outcomes. Frequency and percentages were used for categorical variables; mean (SD) and median (IQR) were used for the continuous variables. Confidence intervals for the intubation and mortality rates, and virologic clearance at day+10 and+30 was calculated using exact binomial. Kaplan-Meier method was used for overall survival (OS). Log-rank statistics was used to compare the OS between product types, donor titers, and pre-treatment immunity. Cox proportional hazards model was utilized to assess the effect of infused plasma neutralizing titers on OS. Univariate test was performed to explore associations between exploratory outcomes and interested groups. Fisher’s exact test was used for categorical variables, and t-test/ANOVA, or its non-parametric version, for the continuous variables based on the normalized of the data. P-value less than 0.05 was considered significant. All statistical analyses were performed using SAS (Version 9.4) and RStudio (Version 0.99.902).

### Study registration

The study is registered with ClinicalTrials.gov NTC04343755, FDA IND approval obtained 4/4/2020 and approved by our Institutional Review Board.

### Patient and public involvement

This study involved the direct participation of the public, as it required volunteer donors. Outreach was done through word of mouth and local media. The public was directed to an institutional webpage with contents approved by our Institutional Review Board.

## Results

Between April 15 and June 16, 2020, 48 patients were enrolled, one had a negative SARS-CoV-2 nasopharyngeal swab RT-PCR and was ineligible. Forty-seven patients were treated, 32 met criteria for track 2, and 15 patients met criteria for track 3. All 47 patients had radiographic evidence of pneumonia. A significant proportion of patients in track 2 were either immunocompromised (22%) or had active cancer (19%), as our hospital harbors the largest cancer center and stem cell transplant program in the State of New Jersey. Demographic and baseline characteristics of patients in track 2 and 3 are summarized in table 1.

**Table 1.** Baseline characteristics of patients with SARS-CoV-2 pneumonia at the time of treatment.

|  | Track 2 (n=32) | Track 3 (n=15) |
| --- | --- | --- |
| **Demographics** |  |  |
| Male sex | 12/32 (62.5%) | 10/15 (67%) |
| Female sex | 20/32 (37.5%) | 5/15 (33%) |
| Median age, years | 59 (IQR 47-69) | 53 (IQR 45-58) |
| Race |  |  |
| Hispanic | 19/32 (59%) | 10/15 (67%) |
| Caucasian | 6/32 (19%) | 3/15 (20%) |
| Black | 1/32 (3%) | 1/15 (7%) |
| Asian | 1/32 (3%) | 1/15 (7%) |
| **Clinical characteristics** |  |  |
| BMI, median | 29 (24-34) | 29 (25-32) |
| Obese or morbidly obese | 14/32 (44%) | 4/15 (27%) |
| Overweight | 7/32 (22%) | 6/15 (40%) |
| Pregnant | 3/32 (9%) | 0 |
| Hypertension | 15/32 (47%) | 6/15 (40%) |
| Diabetes | 9/32 (28%) | 7/15 (47%) |
| Smoking | 4/32 (12.5%) | 2/15 (13%) |
| COPD or asthma | 8/25 (25%) | 1/15 (7%) |
| Immunocompromised | 7/32 (22%) | 1/15 (7%) |
| Active cancer | 6/32 (19%) | 1/15 (7%) |
| **Disease status on day of treatment** |  |  |
| Pneumonia by CXR | 32/32 (100%) | 15/15 (100%) |
| Oxygen supplementation | 29/32 (91%) | 15/15 (100%) |
| 100% non-rebreather mask | 7/32 (22%) | n/a |
| Positive pressure non-invasive mechanical ventilation | n/a | 12/15 (80%) |
| Invasive mechanical ventilation |  | 3/15 (20%) |
| Median days from symptom onset to treatment | 8 (IQR 4-12) | 15 (IQR 10-19) |
| Median days from diagnosis to treatment | 3 (IQR 1-5) | 5 (IQR 4-14) |
| **Concomitant medications** |  |  |
| Hydroxychloroquine | 19/32 (59%) | 12/15 (80%) |
| Azithromycin | 19/32 (59%) | 10/15 (67%) |
| Steroids | 18/32 (56%) | 13/15 (87%) |
| Tocilizumab | 5/32 (16%) | 7/15 (47%) |
| Remdesivir | 4/32 (12.5%) | 2/15 (13%) |
| **Inflammatory markers** |  |  |
| Median ferritin ng/mL | 542 (IQR 200-1160) n=29 | 1520 (IQR 1100-2097) n=13 |
| Median CRP mg/L | 10.6 (IQR 4.3-16.7) n=31 | 3.8 (IQR 1.8-9) |
| Median IL-6 pg/mL | 5 (IQR 5-10) n=15 | 10.5 (IQR 5.5-21.5) |

Among the 32 patients in track 2, 24 (75%) were infused with 500 mL of liquid fresh irradiated plasma and 8 (25%) received 400 mL of fresh frozen plasma. The median dose of plasma IgG_1-4_ infused was 27,537 ug/kg (IQR 21,550-61,408; n=23); 10/32 (31%) received plasma with viral neutralizing titers >1:10,000 and 20/32 (62.5%) with titers 1:1000-10,000. The primary endpoint analysis for track 2 showed that patients had an intubation rate of 15.6% (95% CI: 5.3%-32.8%), this is enough evidence to reject the null hypothesis (p=0.038). Univariate analysis of numerous parameters was performed and is described in table 2. Older age was highly associated with an increased risk of intubation. The false discovery rate (FDR) adjusted p-value for the donor antibody level was not significant. The univariate analysis significance of tocilizumab cannot be ascertained as it was administered to patients with more severe disease.

**Table 2.** Distribution of variables and univariate analysis for patients not mechanically ventilated (track 2)

|  |  | No intubation | Intubation | p-value |
| --- | --- | --- | --- | --- |
| N |  | 27 | 5 |  |
| Age, median |  | 56.00 (IQR 45.00, 65.50) | 69.00 (IQR 66.00, 74.00) | 0.009 |
| Non-rebreather mask O<sub>2</sub>, day 0 | No | 22 (81.5%) | 3 (60.0) | 0.296 |
|  | Yes | 5 (18.5%) | 2 (40.0) |  |
| Hydroxyquinine | No | 12 (44.4%) | 1 (20.0) | 0.625 |
|  | Yes | 15 (55.6%) | 4 (80.0) |  |
| Azithromycin | No | 11 (40.7%) | 2 (40.0) | 1 |
|  | Yes | 16 (59.3%) | 3 (60.0) |  |
| Steroids | No | 12 (44.4%) | 2 (40.0) | 1 |
|  | Yes | 15 (55.6%) | 3 (60.0) |  |
| Tocilizumab | No | 25 (92.6%) | 2 (40.0) | 0.018 |
|  | Yes | 2 (7.4%) | 3 (60.0) |  |
| Remdesivir | No | 23 (85.2%) | 5 (100.0) | 1 |
|  | Yes | 4 (14.8%) | 0 (0.0) |  |
| Product: fresh/frozen | Fresh | 21 (77.8%) | 3 (60.0) | 0.578 |
|  | Frozen | 6 (22.2%) | 2 (40.0) |  |
| Infusion volume, mean |  | 481.48 (SD 39.58) | 440.00 (SD 54.77) | 0.057 |
| Product volume | 400 mL | 5 (18.5%) | 3 (60.0) | 0.085 |
|  | 500 mL | 22 (81.5%) | 2 (40.0) |  |
| Donor titers | > 1:10,000 | 6 (22.2%) | 4 (80.0%) | 0.044 |
|  | 1:1000-10,000 | 19 (70.4%) | 1 (20.0%) |  |
|  | 1:500-1000 | 2 (7.4%) | 0 (0.0%) |  |
| IgM ug/kg infused, median |  | 4678.29 (IQR 3600.38, 6775.45) | 5805.14 (IQR 4742.82, 9295.23) | 0.598 |
| IgG1 ug/kg infused, median |  | 12275.00 (IQR 9514.16, 16289.20) | 12700.30 (IQR 9218.29, 17697.79) | 0.891 |
| IgG2 ug/kg infused, median |  | 9752.68 (IQR 5366.54, 14753.82) | 8300.72 (IQR 6305.89, 10613.50) | 0.441 |
| IgG3 ug/kg infused, median |  | 2428.00 (IQR 1179.64, 4658.96) | 3448.72 (IQR 2754.16, 4300.71) | 0.441 |
| Ig G4 ug/kg infused, median |  | 2415.33 (IQR 891.34, 19128.82) | 18831.48 (IQR 744.27, 41669.43) | 0.968 |
| IgA ug/kg infused, median |  | 5704.39 (IQR 3445.60, 8619.27) | 7800.18 (IQR 533.27, 10360.31) | 0.303 |
| Total IgG ug/kg infused, median |  | 27537.37 (IQR 21550.06, 47961.37) | 47390.46 (IQR 22674.97, 74738.30) | 0.839 |
| Immunity Pre | immunity | 15 (55.6%) | 3 (60.0%) | 1 |
|  | No/minimal | 12 (44.4%) | 2 (40.0%) |  |
| Recipient IgG titers Pre | >1:10,000 | 4 (14.8%) | 0 | 0.946 |
|  | 1:1000-10,000 | 8 (29.6%) | 3 (60%) |  |
|  | 1:500-1000 | 3 (11.1%) | 0 |  |
|  | 1:100-500 | 6 (22.2%) | 1 (20%) |  |
|  | BLQ | 6 (22.2%) | 1 (20%) |  |
| Recipient IgG titers Day3 | >1:10,000 | 8 (33.3%) | 0 | 0.308 |
|  | 1:1000-10,000 | 11 (45.8%) | 2 (50%) |  |
|  | 1:500-1000 | 1 (4.2%) | 1 (25%) |  |
|  | 1:100-500 | 4 (16.7%) | 1 (25%) |  |
| Recipient IgG titers Day10 | > 1:10,000 | 19 (73.1%) | 3 (75%) | 0.508 |
|  | 1:1000-10,000 | 4 (15.4%) | 0 |  |
|  | 1:500-1000 | 1 (12.5%) | 0 |  |
|  | 1:100-500 | 2 (7.7%) | 1 (25%) |  |
|  | BLQ | 1 (3.8%) | 0 |  |
| Recipient titers IgM Pre | Negative | 13 (48.1%) | 2 (40.0) | 1 |
|  | Positive | 14 (51.9%) | 3 (60.0) |  |
| Recipient titers IgM Day 3 | Negative | 2 (8.3%) | 1 (25.0) | 0.382 |
|  | Positive | 22 (91.7%) | 3 (75.0) |  |
| Recipient titers IgM Day 10 | Negative | 3 (11.5%) | 1 (25.0) | 0.454 |
|  | Positive | 23 (88.5%) | 3 (75.0) |  |
| Ferritin Day 0, ng/mL median |  | 525.15 (IQR 198.05, 917.97) n=24 | 792.89 (IQR 763.50, 1972.29) n=5 | 0.312 |
| Ferritin Day 3, median |  | 522.60 (IQR 223.25, 725.26) n=24 | 1496.84 (IQR 1440.53, 1936.41) n=4 | 0.027 |
| CRP Day 0, mg/L median |  | 9.18 (IQR 4.11, 17.00) n=24 | 15.63 (IQR 15.00, 15.98) n=5 | 0.155 |
| CRP Day 3, median |  | 3.25 (IQR 1.93, 9.98) n=23 | 4.10 (IQR 25.35, 35.77) n=4 | 0.015 |
| IL-6 Day 0, pg/mL median |  | 5.00 (IQR 5.00, 10.50) n=14 | 5.00 n=1 | 0.616 |
| IL-6 Day 3, median |  | 5.00 (IQR 5.00, 5.00) n=9 | 203.00 (IQR 107.50, 298.50) n=2 | 0.064 |
| D-dimer Day 0, mg/L median |  | 1.13 (IQR 0.87, 1.55) n=25 | 1.77 (IQR 1.55, 2.12) n=4 | 0.058 |
| D-dimer Day 3, median |  | 1.08 (IQR 0.57, 1.80) n=21 | 2.76 (IQR 1.78, 7.72) n=4 | 0.043 |

Among the 15 patients in track 3, 12 (80%) were infused with 500 mL of liquid fresh irradiated plasma, 3 patients received fresh frozen plasma either 200 mL (1 patient) or 400 mL. The median dose of infused plasma IgG_1-4_ ug/kg was 38,260 (IQR 33,3076-50,426; n=12); 5/15 (33.3%) received plasma with neutralizing titers >1:10,000 and 9/15 (60%) with titers 1:1000-10,000. The primary endpoint analysis for track 3 showed that patients had a day-30 mortality of 46.7% (7/15; 95% CI:21.3%-73.4%). This track 3 mortality rate was compared to institutional data of patients with SARS-CoV-2 pneumonia, in ICU, receiving invasive or non-invasive mechanical ventilation. 317 patients met these criteria with a mortality rate of 68.5% (217/317; p=0.093). Track 2 and 3 survival comparison with institutional data is shown in table 4. Univariate analysis of numerous parameters was performed and is described in table 3. The overall survival plots for each track is shown in figures 1 and 2. There was a single adverse event for all 47 patients, one patient developed a grade 2 rash (CTCAE v4.0) for which hydrocortisone 100 mg IV was administered once with resolution.

**Table 3.** Distribution of variables and univariate analysis for patients on positive pressure mechanical ventilation (track 3).

|  |  | Alive | Dead | p-value |
| --- | --- | --- | --- | --- |
| N |  | 8 | 7 |  |
| Age, median |  | 55.50 (IQR 50.00, 58.75) | 49.00 (IQR 41.00, 56.00) | 0.267 |
| Hydroxyquinine | No | 2 (25%) | 1 (14.3%) | 1 |
|  | Yes | 6 (75%) | 6 (85.7%) |  |
| Azithromycin | No | 4 (50%) | 1 (14.3%) | 0.282 |
|  | Yes | 4 (50%) | 6 (85.7%) |  |
| Steroids | No | 2 (25%) |  | 0.467 |
|  | Yes | 6 (75%) | 7 (100%) |  |
| Tocilizumab | No | 5 (62.5%) | 3 (42.9%) | 0.619 |
|  | Yes | 3 (37.5%) | 4 (57.1%) |  |
| Remdesivir | No | 6 (75%) | 7 (100%) | 0.467 |
|  | Yes | 2 (25%) | 0 |  |
| Product: fresh/frozen | Fresh | 6 (75%) | 6 (85.7%) | 1 |
|  | Frozen | 2 (25%) | 1 (14.3%) |  |
| Infusion volume, mean |  | 450.00 (SD 106.9) | 485.71 (SD 37.8) | 0.619 |
| Donor IgG titers | >1:10,000 | 2 (25%) | 3 (42.9%) | 1 |
|  | 1:1000-10,000 | 5 (62.5%) | 4 (57.1%) |  |
|  | 1:500-1000 | 1 (12.5%) | 0 |  |
| IgM ug/kg infused, median |  | 6638.97 (IQR 5439.39, 8279.42) | 4475.55 (IQR 3920.08, 6727.85) | 0.194 |
| IgG1 ug/kg infused, median |  | 16535.29 (IQR 12813.00, 22444.04) | 12009.06 (IQR 10317.84, 15954.94) | 0.256 |
| IgG2 ug/kg infused, median |  | 15094.85 (IQR 8622.97, 18774.82) | 7389.73 (IQR 6058.87, 12827.95) | 0.104 |
| IgG3 ug/kg infused, median |  | 3150.47 (IQR 3031.66, 3281.62) | 4169.17 (IQR 2343.29, 9080.77) | 0.745 |
| IgG4 ug/kg infused, median |  | 4931.74 (IQR 1312.22, 13930.51) | 1513.87 (IQR 657.88, 5799.86) | 0.626 |
| IgA ug/kg infused, median |  | 6379.23 (IQR 6377.62, 10133.75) | 6564.96 (IQR 5763.48, 6774.62) | 0.871 |
| Total IgG ug/kg infused, median |  | 46536.05 (IQR 35410.90, 73192.06) | 33628.45 (IQR 29786.79, 43242.17) | 0.104 |
| Recipient IgG titers Pre | >1:10,000 | 2 (25%) | 2 (28.6%) | 0.765 |
|  | 1:1000-10,000 | 6 (75%) | 4 (57.1%) |  |
|  | 1:500-1000 | 0 | 1 (14.3%) |  |
| Recipient IgG titers Day 3 | >1:10,000 | 6 (75%) | 6 (85.7%) | 1 |
|  | 1:1000-10,000 | 2 (25%) | 1 (14.3%) |  |
| Recipient IgG titers Day 10 | >1:10,000 | 7 (87%) | 4 (80%) | 1 |
|  | 1:1000-10,000 | 1 (12.5%) | 1 (20%) |  |
| Ferritin Day 0, ng/mL median |  | 1778.07 (IQR 1057.64, 2109.22) n=8 | 1288.90 (IQR 1186.70, 1635.94) n=5 | 0.942 |
| Ferritin Day 3, median |  | 1046.03 (IQR 745.52, 2187.83) n=7 | 1448.58 (IQR 999.67, 2370.05) n=6 | 0.830 |
| Ferritin Day 10, median |  | 1102.88 (IQR 956.82, 1248.94) n=2 | 1479.57 (IQR 1098.72, 1642.57) n=5 | 0.561 |
| CRP Day 0, mg/L median |  | 2.34 (IQR 1.73, 10.82) n=8 | 6.86 (IQR 3.17, 8.86) n=7 | 0.524 |
| CRP Day 3, median |  | 6.1 (IQR .28, 14.2) n=7 | 7.19 (IQR 2.62, 16.89) n=7 | 0.443 |
| CRP Day 10, median |  | 2.27 (IQR 1.57, 3.96) n=3 | 7.12 (IQR 3.57, 14.98) n=4 | 0.596 |
| IL-6 Day 0, pg/mL median |  | 13.00 (IQR 11.25, 25.50) n=4 | 6.00 (IQR 5.00, 19.75) n=6 | 0.331 |
| IL-6 Day 3, median |  | 9.00 (IQR 4.75, 29.50) n=3 | 9.00 (IQR 5.00, 75.25) n=6 | 0.694 |
| IL-6 Day 10, median |  | 5.00 n=1 | 79.00 (IQR 51.00, 212.50) n=3 | 0.371 |
| D-dimer Day 0, mg/L median |  | 1.84 (IQR 1.59, 11.15) n=7 | 2.65 (IQR 1.58, 6.29) n=7 | 0.848 |
| D-dimer Day 3, median |  | 2.17 (IQR 1.35, 12.50) n=7 | 5.89 (IQR 3.56, 11.64) n=7 | 0.522 |
| D-dimer Day 10, median |  | 1.92 (IQR 1.39, 5.75) n=3 | 2.35 (IQR 1.43, 5.76) n=5 | 0.766 |

**Table 4.** Survival comparison of track 2 and 3 with institutional control.

|  |  | Track 2 | Track 2 control | p-value |
| --- | --- | --- | --- | --- |
|  | n | 32 | 1023 |  |
| Survival status (%) | alive | 28 (87.5%) | 675 (66%) | 0.012 |
|  | dead | 4 (12.5%) | 348 (34%) |  |
|  |  | Track 3 | Track 3 control | p-value |
|  | n | 15 | 317 |  |
| Survival status (%) | alive | 8 (53.3%) | 100 (31.5%) | 0.093 |
|  | dead | 7 (46.7%) | 217 (68.5%) |  |

**Figure 1.**
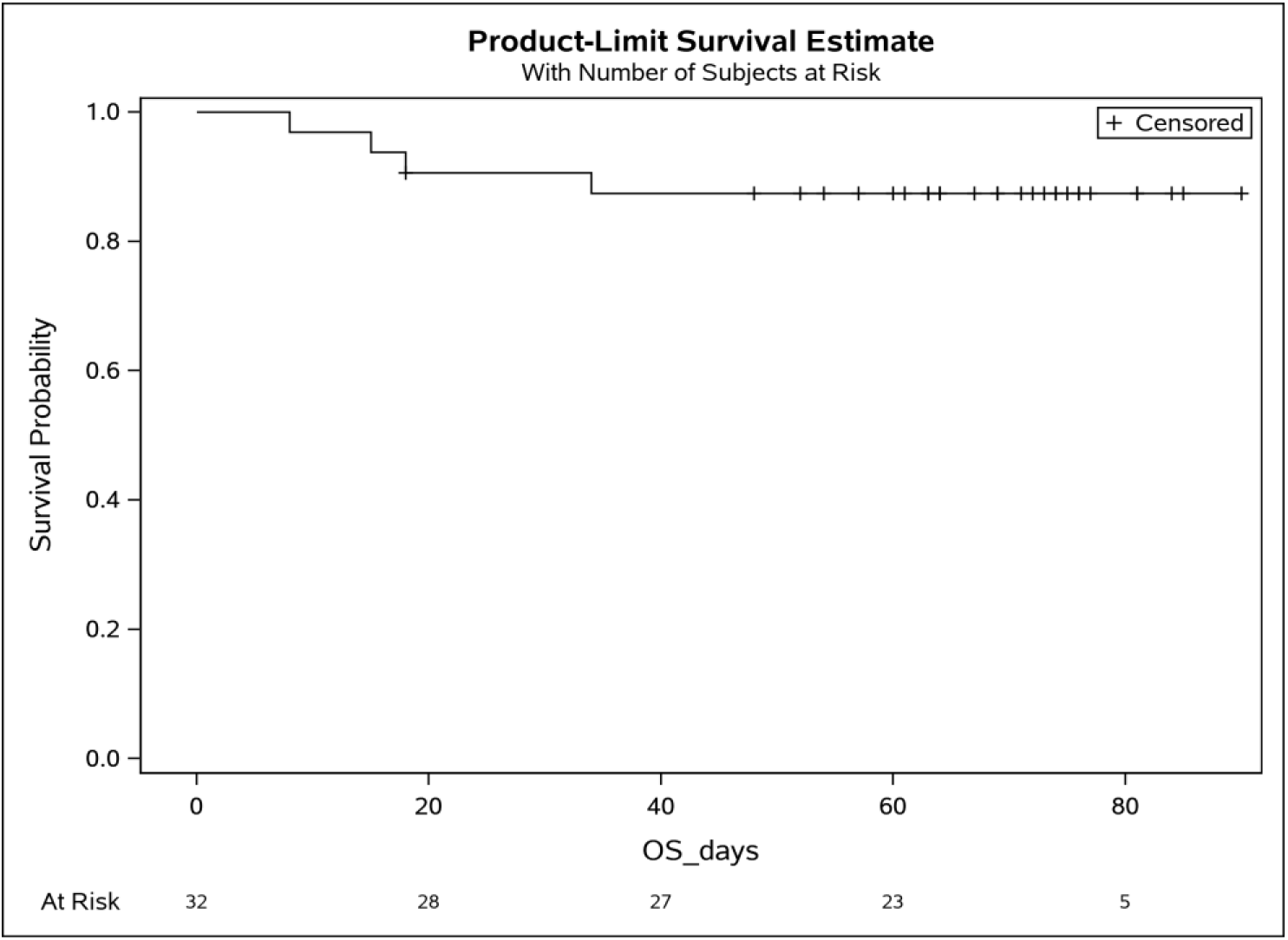
Overall survival for patients non-mechanically ventilated (track 2)

**Figure 2.**
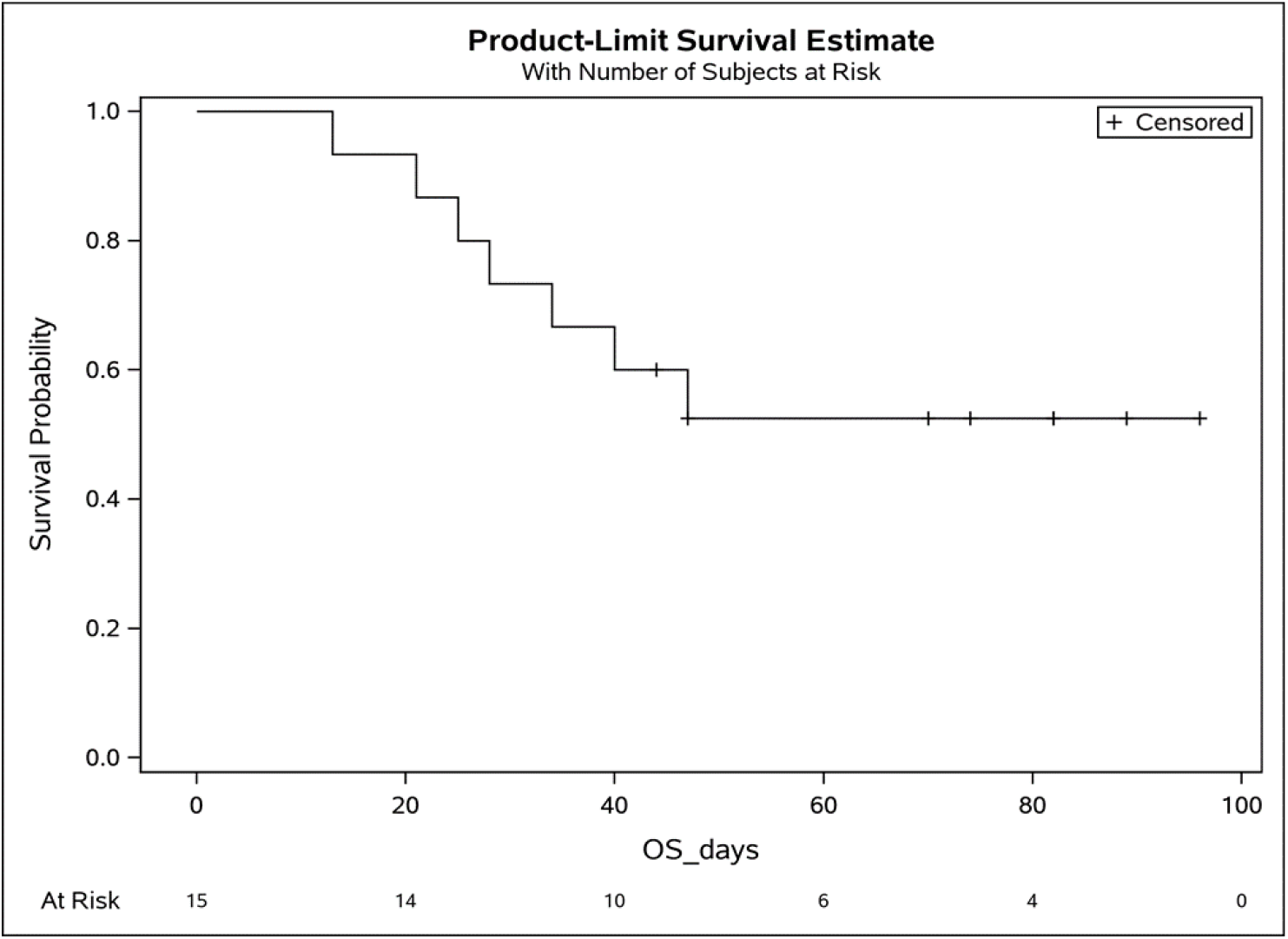
Survival of patients on positive pressure mechanical ventilation (track 3)

Secondary endpoints analysis for track 2 demonstrated a day-30 survival rate of 87.5% (28/32; 95% CI: 70.2%-96.4%). This was compared with institutional survival data for patients with SARS-CoV-2 pneumonia receiving oxygen, excluding patients receiving positive pressure mechanical ventilation. 1023 patients met these criteria with a survival rate of 66% (675/1023; p=0.012), described in table 4. The rates of negative nasopharyngeal swab by PR-PCR at day+10 and +30 post treatment were 42.9% (95% CI: 24%-63%; n=28) and 78% (95% CI: 56%-63%; n=23) respectively, in the context of a median time from symptom onset to treatment of 8 days (IQR 4-12). There was only one COVID-19-related readmission and the patient was subsequently discharged. Secondary endpoints analysis for track 3 demonstrated rates of negative nasopharyngeal swab or endotracheal secretion analysis by PR-PCR at day+10 and +30 of 85.7% (95% CI: 42-100%; n=7) and 100% (95% CI: 63-100%; n=8) respectively, with a median time from symptom onset to treatment of 15 days (IQR 9-19). The day-30 discharged alive rate was 46.7% (7/15) with one patient extubated but not yet discharged. There were no readmissions.

For either tracks, there was no statistically significant difference in survival, duration of hospitalization, post infusion antiviral titers, and post infusion inflammatory markers (CRP, ferritin, IL-6 and D-dimers) between fresh and frozen plasma, infused plasma immune globulin subtype (IgA, IgM, IgG_1-4_) content, or concomitant medications (listed in table 1). There was also no difference in these endpoints within the ranges of donor IgG antiviral titers used, which were all above >1:500 (2 donors) and predominantly >1:1000 (tables 2 and 3).

Transfer of immunity was evaluated by measuring the recipients’ anti-SARS-CoV-2 neutralizing titer levels immediately pre-infusion and again on day+3. Seven patients (15%), all in track 2 had no pre-infusion titers, and subsequently all 7 were found to have anti-SARS-CoV-2 neutralizing titers on day+3. One transplant patient on immunosuppression was found to have undetectable titers on day+10. Patients in track 3 all had anti-SARS-CoV-2 titers pre-infusion, 4/15 (27%) >1:10,000, 10/15 (67%) 1:1000-10,000, and 1/15 (7%) 1:500-1000. However, we observed an increase on day+3 with 12/15 (80%) >1:10,000 and 3/15 (20%) 1:1000-10,000. All evaluated patients on study were found to have neutralizing titers on day+30 (n= 30) and on day+60 (n=12) (figure 3).

**Figure 3.**
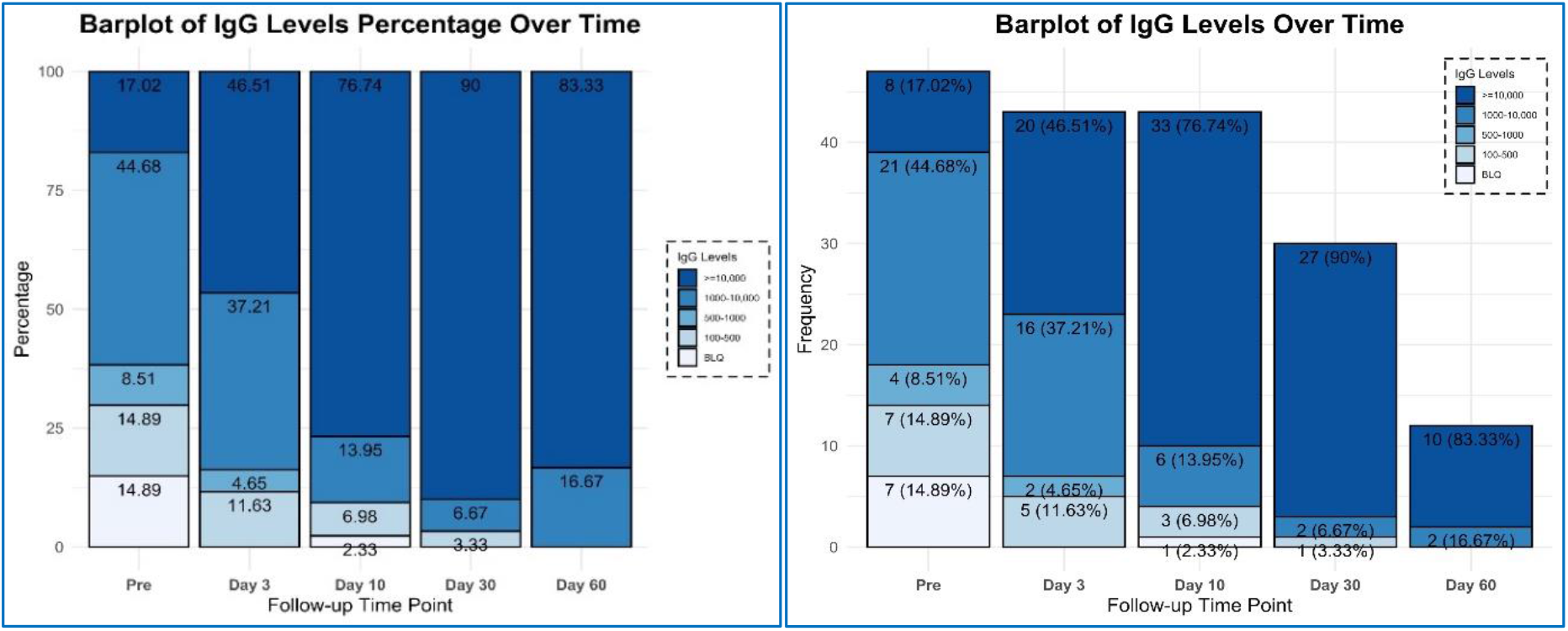
Recipients neutralizing antibody titers percentage and frequency over time for all patients.

We performed a post hoc analysis of the patients who received convalescent plasma while either non-immune or minimally immune defined as titers 1:100-500. Fourteen patients met these criteria, all in track 2. The intubation rate was 14.3% (95% CI: 1.8%-42.8%). Day+10 and +30 viral clearance by nasopharyngeal swab was 50% (95% CI: 21%-79%) and 78% (95% CI 40-98%) respectively. Immunity over time is shown in figure 4. There was a trend but no statistically significant difference in overall survival between the non- or minimally immune and the immune group (figure 5). Of note, the only 2 deaths in the non-immune group were attributed to patients with active lymphoma on chemotherapy, one peripheral T cell lymphoma and one with refractory relapsed chronic lymphocytic leukemia.

**Figure 4.**
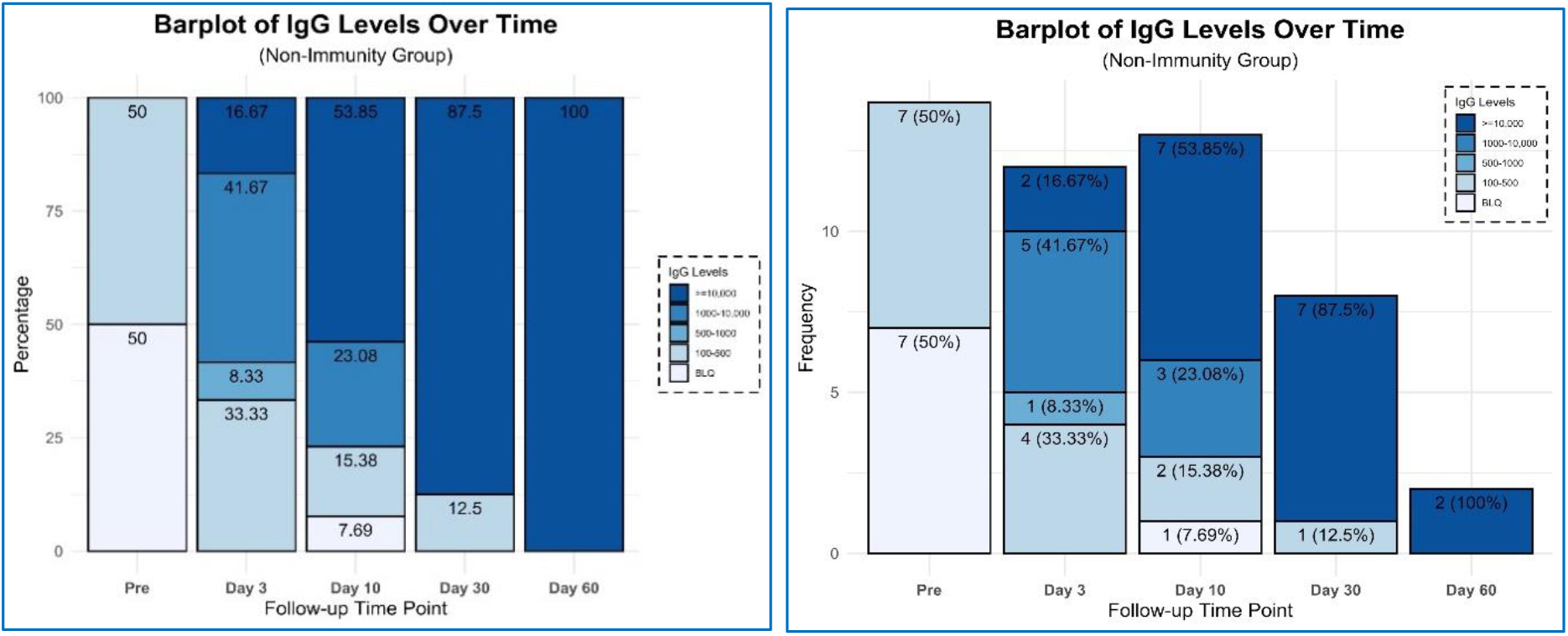
Neutralizing antibody titers percentage and frequency over time for non-immune or minimally immune patients.

**Figure 5.**
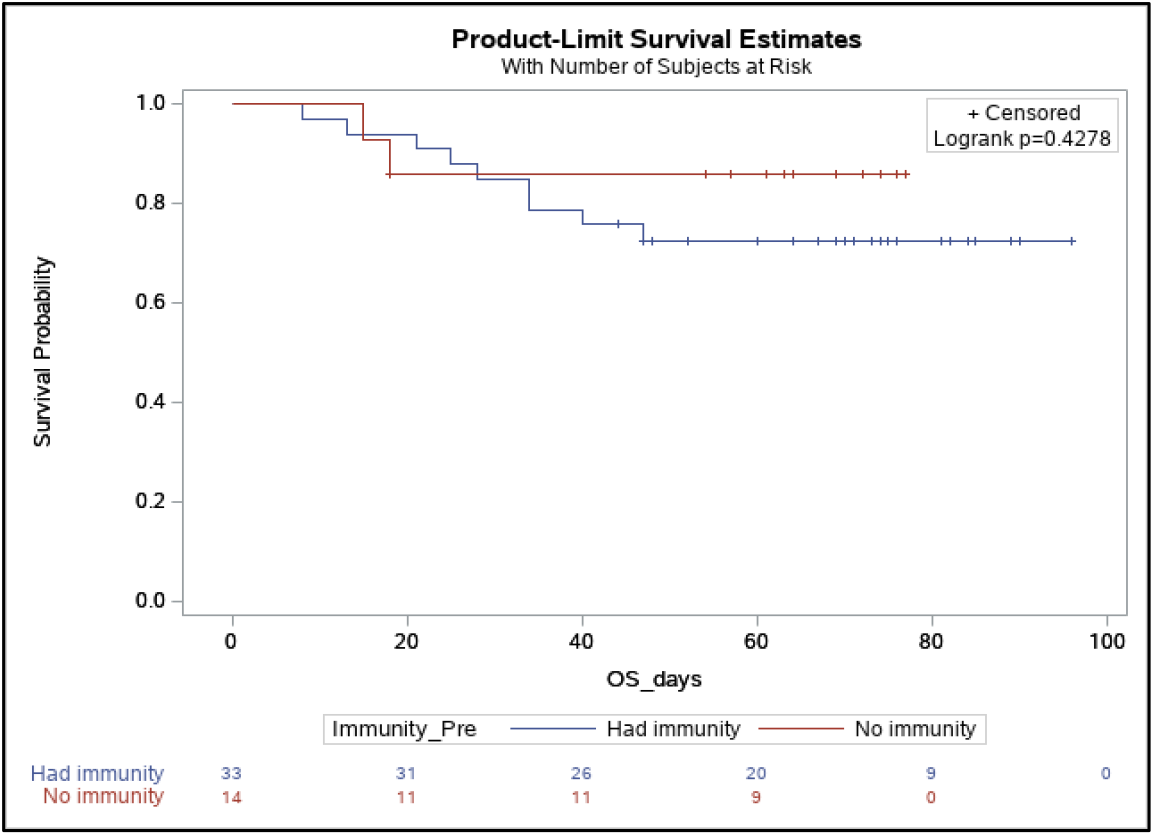
OS immune vs non/minimally immune group.

## Discussion

### Principle Findings

In this prospective study investigating the therapeutic use of convalescent plasma in patients with COVID-19 disease, we showed that the administration of high-titer donor plasma is safe and effectively transfers antiviral titers, while preserving the endogenous development of immunity. The study was conducted at the height of the epidemic in New Jersey, when most patients were hospitalized only if requiring oxygen supplementation. In congruence with this fact, all patients treated had pneumonia. Only 13% of patients concomitantly received remdesivir, allowing for the evaluation of convalescent plasma as the sole antiviral agent administered for most patients. Our results showed an intubation rate of 15.6% and for the ventilated patients a day-30 mortality of 46.7%. The overall survival of the treated patients compares favorably with our institutional database, reaching statistical significance for non-mechanically ventilated patients. Within the ranges of plasma antiviral titers above 1:1000, we were not able to see a difference in outcome based on titer levels. Frozen plasma was not inferior to fresh plasma. Plasma was infused without adverse events, except for one mild rash, to a wide spectrum of recipients including those who were ventilated, elderly, pregnant and immunocompromised.

### Comparison with other studies

In the search for antiviral therapy, our findings clearly demonstrate the safety of convalescent plasma and the passive immunity transfer. As the original data from China used fresh liquid plasma^11^ and most centers in the United States make use of fresh frozen plasma, the lack of a significant difference between these products is important information. Frozen plasma allows the flexibility of use, as it can be accumulated and rapidly deployed during a viral surge. Since most of the plasma were from donors with titers above 1:1000, we cannot determine a lowest level acceptable. However, we can ascertain within the statistical limits of this study that we need not limit our donor pool to those with the higher titers >1:10,000, and a cut-off of 1:1000 will be used for our subsequent studies.

Early viral neutralization, with the ensuing prevention of the catastrophic immune response to viral damage, forms the basis for the infusion of high-titer convalescent plasma. Our expectation at protocol inception was to have access to patients early in the course of their disease. The reality, however, of conducting a clinical trial in the setting of an overwhelming influx of cases meant that most patients were not hospitalized until later in their course, during the inflammatory phase. We therefore conducted an ad hoc analysis of the non-immune patients which included patients early in their course and patients unable to mount an immunity, such as immunocompromised patients. Understanding the kinetics of immune response to the virus is important and has been recently elegantly described. In a series of 23 patients with mild or severe disease,^26^ IgG antibodies emerged at 10-15 days post onset of symptoms, were sustained for at least 6 weeks and with a similar IgG response for both the mild and severe groups. Based on these kinetic descriptions, we can confirm that the presence of antibodies on day+3 was from passive transfer and not time related. Interestingly, the same authors reported that most patients with severe disease still had viral shedding 30-40 days post onset of disease, bringing into question the neutralizing capability of those endogenous antibodies.^27^ In our study, recipients demonstrated a high level of viral clearance at post infusion day+10 and +30.

Track 3 represents a group of severely ill patients, either non-invasively or invasively ventilated, all with endogenous immune titers. Our management of patients with COVID-19 reserved invasive ventilation almost exclusively for patients failing non-invasive positive pressure ventilation measures. The clinical course and outcomes of critically ill patients with SARS-CoV-2 pneumonia has been previously reported.^28,29^ In a series of 52 patients similar to our track 3 patients, receiving either invasive or non-invasive mechanical ventilation, 32 (61.5%) had died by day 28, and of the remaining 20 patients, only 8 (15.4%) were discharged.^28^ In our current study, track 3 patients had a day-30 discharge alive rate of 46.7% and a viral clearance of 86.7% at day+10 post treatment. This may support the position that passive transfer of antiviral titers may be of benefit even in patients with immunity.

The focus of most antiviral therapy has been early in the course of the disease. In comparison to patients in track 3, patients in track 2 had a shorter time from symptoms onset to treatment. The track 2 day-30 discharge alive rate was 87.5%, even though 22% of patients were immunocompromised either from cancer or transplantation, 100% had pneumonia, and 91% required oxygen supplementation. The performance of the non-immune group is of most interest, as the only fatalities in this group were attributed to the two patients with advanced lymphoma on active chemotherapy. Our day-30 discharge alive rate for this patient population was 85.7% (12/14). A recent randomized study evaluated the effect of convalescent plasma on the time to symptom improvement in severe COVID-19 disease.^11^ Patients were excluded if they had high titers of S-protein-RBD-specific IgG antibodies (≥ 1:640), leaving a similar patient population to our non-immune or minimally immune patients. The median volume infused was 200 mL compared to 400-500 mL in our study. In this randomized study the day-28 mortality was 15.7% for the patients in the plasma group, with a discharge rate of 51%. Details of the plasma content or immunity transfer was not provided. There was a statistically significant increase in the rate of viral negativity by PCR in the plasma group, but no difference in the primary outcome of time to clinical improvement. This study was unfortunately limited by the small sample size.

### Limitation of this study

Our study was limited by the lack of randomization to a control group, and the access to patients early in the disease course, where antiviral interventions is presumed to be of greatest impact. Our study was also not powered or designed to evaluate the optimal donor antiviral titer level, or the optimal dose of IgM and IgG to be infused. We are conducting a randomized study of convalescent plasma in high-risk patients with early onset disease with the aim of reducing hospitalizations.

## Conclusion

In conclusion, we aimed at gaining a better understanding of the clinical and laboratory effects of high-titer convalescent plasma in hospitalized patients with severe COVID-19 pneumonia. We found that the infusion of convalescent plasma is safe, effectively transfers of antiviral immunity, leads to a high incidence of viral clearance, and does not preclude the development of endogenous immunity. The low rate of intubation and the survival at day 30 are encouraging and warrant further evaluation within the context of a randomized study.

We acknowledge the assistance of Professor Florian Krammer PhD, Krammer Laboratory, Icahn School of Medicine at Mount Sinai, New York, USA, in the development and early reporting of the Spike-RBD ELISA, and in making available for us the recombinant proteins. We also acknowledge the help and support of Drs. Ihor Sawczuk and Lisa Tank in their leadership roles at Hackensack University Medical Center. We extend our deepest gratitude to the hundreds of volunteer donors who reached out to participate in this study.

## Data Availability

In Manuscript

